# Endovascular Robotic-Assisted SystEm for Cerebral Angiography (ERASE): Rationale, Design, and Protocol of a Multicenter Randomized Controlled Trial

**DOI:** 10.64898/2025.12.19.25342721

**Authors:** Yuanli Zhao, Siming Gui, Jia Jiang, Yang Zhao, Qi Liu, Jianbo Chang, Anqi Li, Xiaobo Zhang, Fei Wang, Xiaoqing Zhang, Jun Lin, Yuhua Jiang, Xinke Liu, Ya Gao, Xiangyu Meng, Wei You, Wentao Gong, Sheng Guan, Yong Sun, Xiang Xu, Conghui Li, Youxiang Li

**Affiliations:** Department of Neurosurgery, Peking Union Medical College Hospital, Peking Union Medical College, Chinese Academy of Medical Sciences, Beijing, China; Department of Interventional Neuroradiology, Beijing Neurosurgical Institute, Capital Medical University, Beijing, China; Department of Neurosurgery, Beijing Tiantan Hospital, China National Clinical Research Center for Neurological Diseases, Capital Medical University, Beijing, China; Department of Radiology, Peking Union Medical College Hospital, Peking Union Medical College, Chinese Academy of Medical Sciences, Beijing, China; Department of Operating Room, Peking Union Medical College Hospital, Peking Union Medical College, Chinese Academy of Medical Sciences, Beijing, China; Department of Operating Room, Beijing Tiantan Hospital, China National Clinical Research Center for Neurological Diseases, Capital Medical University, Beijing, China; Department of Neurosurgery, The First Hospital, Hebei Medical University, Shijiazhuang, Hebei, China; Department of Interventional Neuroradiology, The First Affiliated Hospital of Zhengzhou University, Zhengzhou, Henan, China; Department of Neurosurgery, The First People’s Hospital of Lianyungang, Nanjing Medical University, Lianyungang, Jiangsu, China; Department of Neurosurgery, Tangshan Gongren Hospital, Hebei Medical University, Tangshan, Hebei, China

**Author notes:** Correspondence: Prof. Youxiang Li, Department of Neurosurgery, Beijing Tiantan Hospital, China National Clinical, Research Center for Neurological Diseases, Capital Medical University, Beijing, China;, Prof. Conghui Li, Department of Neurosurgery, The First Hospital, Hebei Medical University, Shijiazhuang, Hebei, China;, Prof. Xiang Xu, Department of Neurosurgery, Tangshan Gongren Hospital, Hebei Medical University, Tangshan, Hebei, China. These authors contribute equally to this work (Yuanli Zhao, Siming Gui, Jia Jiang).

## Abstract

**Background:** Neurointerventional therapy is a cornerstone in managing head and neck vascular disorders, with cerebral angiography serving as its fundamental diagnostic and therapeutic backbone. However, manual cerebral angiography is associated with several inherent limitations. While existing robotic-assisted systems have shown promise in mitigating some of these issues, they face challenges such as limited compatibility, lengthy setup times, and a lack of high-quality real-world evidence.

**Methods:** The ERASE trial is a multicenter, prospective, randomized controlled trial (RCT). A total of 450 eligible patients will be enrolled from six comprehensive stroke centers in China and randomized 1:1 to either the robotic-assisted group or the control group. Both groups use the Seldinger technique for femoral/radial artery access. Operators undergo standardized training on the robotic system, and all patients are followed up at baseline, end of surgery, 24 hours postoperatively, and 7 days post-discharge.

**Results:** The primary efficacy outcome is the clinical success rate. The primary safety outcome is the incidence of perioperative/postoperative complications (e.g., vascular perforation, dissection, pseudoaneurysm), serious adverse events, and device malfunctions. Secondary outcomes include technical failure rate, overall procedural time, pre-puncture setup time, target vessel super-selective catheterization time, digital subtraction angiography fluoroscopy time, participant radiation doses and contrast agent volume. A key safety endpoint is the rate of new asymptomatic cerebral infarctions detected via postoperative brain MRI-diffusion-weighted imaging.

**Conclusions:** As the RCT focusing on the YDHB-NS01 Ver 2.0 system, the ERASE trial addresses critical unmet needs in neurointerventional practice and will generate high-quality evidence for robotic-assisted cerebral angiography.

**Trial registration number:** ClinicalTrials.gov NCT07182188.

**Clinical Perspective:**

1) What Is New?

This multicenter RCT evaluates the YDHB-NS01 Ver 2.0 robotic-assisted system and provides rigorous evidence on its safety and efficacy compared with manual cerebral angiography, while validating targeted design enhancements addressing prior systems’ shortcomings

2) What Are the Clinical Implications?

The study’ s findings could standardize the clinical application of robotic-assisted cerebral angiography and inspire further research on refining robotic interventional workflows to improve patient outcomes and provider safety.

## Introduction and rationale

Today, neurointerventional therapy has been widely adopted and plays an increasingly pivotal role in the management of hemorrhagic conditions such as intracranial aneurysms, vascular malformations, and nonacute subdural hematomas as well as ischemic disorders including acute large vessel occlusion ^1–6^. Cerebral angiography stands as a foundational and indispensable component of neurointerventional procedures, serving multiple critical functions: the diagnosis of cerebrovascular diseases, the development of treatment strategies, and postprocedural imaging follow-up ^7–11^.

Robots are electromechanical systems designed to execute repetitive, demanding tasks, or tasks requiring exceptional precision ^7^ ^12^. The core objective of any robotic surgical system is to enhance human capabilities by surpassing the constraints of inherent physical limitations and enabling remote surgical execution. In the context of cerebral angiography, endovascular robotic-assisted platforms aim to improve surgical precision and safety, optimize patient outcomes (including treatment success rates and comfort), minimize variability in operator performance, and reduce radiation exposure for both patients and clinicians. Furthermore, leveraging the remote operation capabilities of these systems, patients in medically underserved areas can gain access to such advanced technologies ^7^ ^13^. Although numerous studies have explored robotic-assisted cerebral angiography and compared it with manual procedures, current robotic systems still suffer from several key limitations: prolonged preoperative setup and installation times, limited compatibility with commercially available neurointerventional devices (e.g., guidewires and catheters) and digital subtraction angiography (DSA) systems from various manufacturers, small sample sizes in existing clinical trials, restriction to transfemoral access, and a predominance of premarket exploratory studies ^14–16^.

In response to these challenges, we have developed a new-generation robotic-assisted system specifically designed for neurointerventional cerebral angiography: the YDHB-NS01 Ver 2.0 (Yidu Hebei Robot Technology Co., Ltd., Hebei, China). This system incorporates numerous design enhancements and functional optimizations and has successfully obtained a medical device registration certificate from China’s National Medical Products Administration (Figure 1). The YDHB-NS01 Ver 2.0 features two delivery devices with clamping mechanisms, enabling simultaneous, independent control of two instruments: one guidewire and one catheter. These mechanisms allow for precise millimeter-scale adjustments to advance, retract, rotate, or perform complex combined movements (e.g., simultaneous advancement and rotation), facilitating accurate delivery of catheters and guidewires. The system is compatible with a wide range of neurointerventional devices, including 5-Fr angiographic catheters, 6-Fr guiding catheters (length up to 125 cm), and 0.035-inch guidewires (length up to 260 cm). Compatible catheter types include Pigtail, VER, Simmons (notably utilized for transradial access), MPA1, and Hunter-Head catheters. Additionally, the system supports long-distance delivery of guidewires and catheters, ranging from the femoral artery to the cavernous segment of the internal carotid artery (ICA). Regarding the physician’s console, it features dedicated control handles for catheter and guidewire delivery. During the procedure, movements of the catheter and guidewire manipulators on the robotic platform are synchronized with those of the handles. When the handles corresponding to the catheter and guidewire are pushed forward or pulled backward, the respective manipulators advance and retract at a fixed speed accordingly. Conversely, rotating the handles clockwise or counterclockwise triggers the manipulators to rotate the catheter or guidewire in the respective direction.

**Figure 1.**
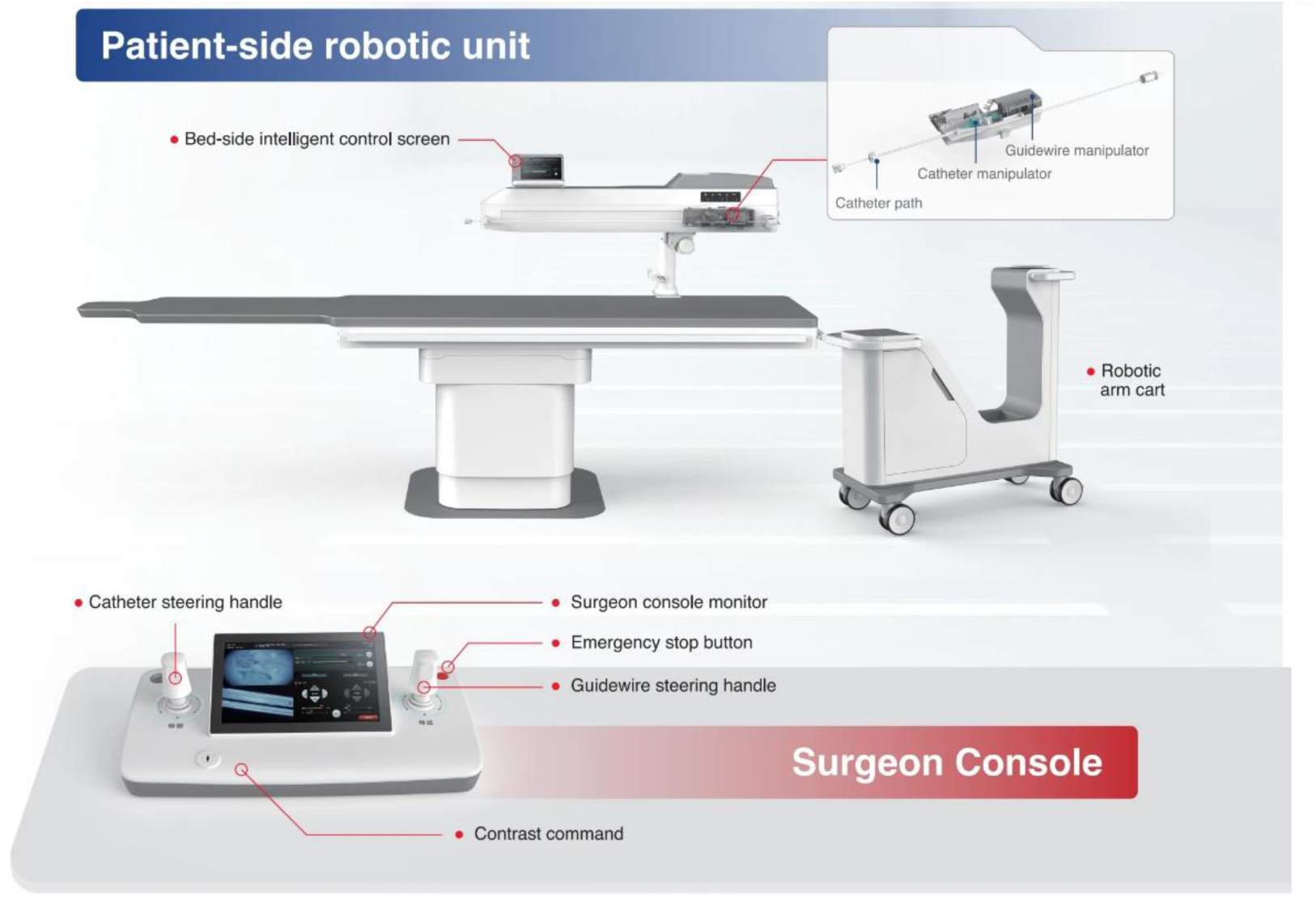
Components of the YDHB-NS01 Ver 2.0 robotic-assisted system. The patient-side robotic unit and the Surgeon-side control console. Both components can be mounted on a portable mobile cart for transportation.

However, high-quality evidence from randomized controlled trials (RCTs) remains limited regarding the safety and efficacy of this robotic-assisted system in cerebral angiography. Therefore, we designed the Endovascular Robotic-Assisted SystEm for Cerebral Angiography (ERASE) RCT to compare robotic-assisted versus manual cerebral angiography, with the aim of generating high evidence supporting the clinical utility of endovascular robotic-assisted systems for this procedure.

## Methods

### Study Design and Patient Population

The ERASE trial is an investigator-initiated, multicenter, prospective, randomized, open-label, blinded-endpoint study designed to enroll 450 patients. Patients with suspected head and neck vascular lesions requiring cerebral angiography for definitive diagnosis, who meet the ERASE trial’s inclusion and exclusion criteria (Table 1), will be considered for enrollment across six comprehensive stroke centers in China (ClinicalTrials.gov identifier: NCT07182188). The study protocol has been approved by the institutional review board (IRB) of each participating center.

**Table 1.** Inclusion and exclusion criteria of ERASE trial.

|  |  |
| --- | --- |
| **Inclusion criteria:** |  |
| (1) | Age 18–80 years; |
| (2) | Suspected head and neck vascular lesions or other conditions involving head and neck vessels that require cerebral angiography for definitive diagnosis or to guide treatment decision-making, including but not limited to: intracranial aneurysms, arteriovenous malformations (AVMs), dural arteriovenous fistulas (DAVFs), Moyamoya disease (MMD), subarachnoid hemorrhage (SAH), symptomatic intracranial artery stenosis/occlusion, carotid/subclavian/vertebral artery stenosis/occlusion, acute or chronic subdural hematoma, and carotid body tumors; |
| (3) | American Society of Anesthesiologists (ASA) Physical Status Class I–III; |
| (4) | Provision of written informed consent by all patients or their legally authorized representatives prior to enrollment. |
| **Exclusion criteria:** |  |
| (1) | Patients with underlying hereditary systemic connective tissue disorders; |
| (2) | Documented allergy to iodinated contrast media; |
| (3) | Severe local infection, skin ulceration, or significant vascular anatomical abnormalities at the planned puncture site; |
| (4) | Inability to undergo brain MRI (e.g., due to non-MRI-compatible metallic implants, claustrophobia, or other established contraindications); |
| (5) | Laboratory or clinical abnormalities including: platelet count $< 90 \times 10^9/L$, hematocrit $< 30\%$ (0.30), international normalized ratio (INR) $> 1.5$, uncontrolled severe hypertension despite medical management (systolic blood pressure [SBP] $> 180$ mmHg or diastolic blood pressure [DBP] $> 110$ mmHg), severe heart or respiratory failure, severe hepatic impairment (alanine transaminase [ALT] or aspartate transaminase [AST] $> 3 \times$ upper limit of normal [ULN]), or renal dysfunction (serum creatinine $> 3$ mg/dL [$> 265 \mu$ mol/L]); |
| (6) | Concurrent participation in another interventional drug or medical device clinical trial; |
| (7) | Pregnancy, lactation, or plans to become pregnant within 1 year (for women of childbearing potential); |
| (8) | Inability to complete study follow-up due to cognitive impairment, emotional disorders, or psychiatric illness; |
| (9) | Life expectancy $< 3$ years; |
| (10) | Other conditions deemed inappropriate for enrollment by the investigating team. |

ERASE, Endovascular Robotic Assisted SystEm for Cerebral Angiography; MRI, magnetic resonance imaging.

A flow chart outlining the ERASE trial is presented in Figure 2. The ERASE trial utilized an interactive web response system (IWRS) for central randomization, stratified by study centers. Following the entry of the pertinent information of qualifying patients into the web-based system, the researchers at each center will obtain a random code as well as the corresponding group allocation information from the IWRS. Eligible patients will be randomized to one of two treatment groups: the robot-assisted group and the control group. Those assigned to the robot-assisted group will undergo cerebral angiography performed by physicians via the YDHB-NS01 Ver 2.0 robotic-assisted system, while those in the control group will undergo the procedure via manual techniques by physicians. To ensure patient safety, physicians are required to have > 1 years of experience in cerebral angiography. All physicians completed dedicated training sessions using silicone vascular models prior to clinical procedures to familiarize them with robotic system assembly and operational steps. The goal of these training sessions was to simulate clinical scenarios, including sterile technique implementation, full angiographic procedural workflows, and emergency management. Physicians practiced selective catheterization of bilateral ICAs and bilateral vertebral arteries (VAs), as well as emergency protocols (e.g., robotic system emergency stop and rapid conversion to manual operation).

**Figure 2.**
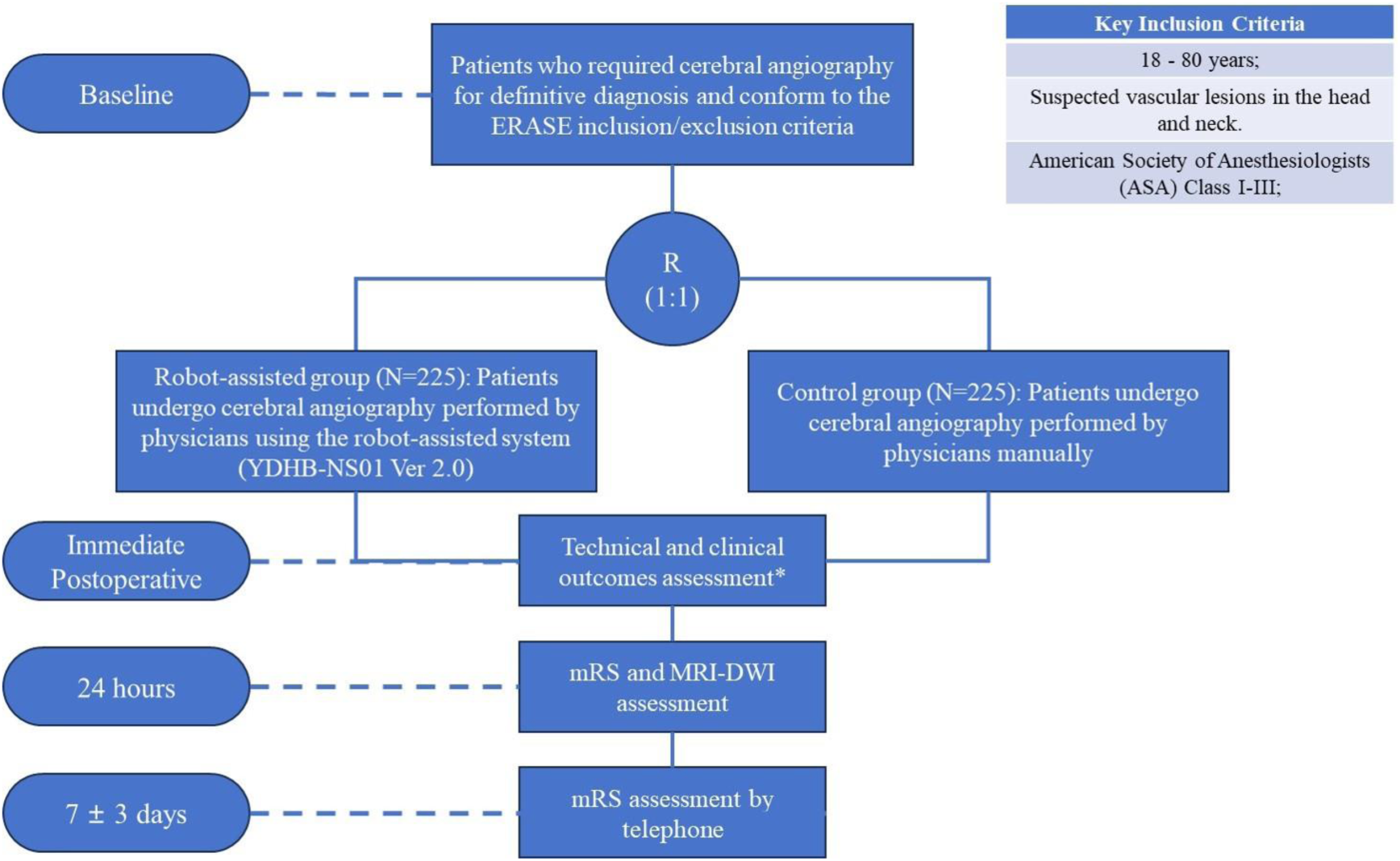
The flowchart of ERASE trial. *Technical and clinical outcomes assessment includes clinical success Rate, super-selective technique success rate, super-selective catheterization time for target vessels, pre-puncture set up time, DSA fluoroscopy time, overall surgical time, participant radiation dose, operator radiation dose, contrast agent dosage and instrument performance evaluation; mRS, modified Rankin Scale; ERASE, Endovascular Robotic Assisted SystEm for Cerebral Angiography; MRI, magnetic resonance imaging; DWI, diffusion-weighted imaging.

### Intervention Procedure

In both the robotic-assisted and manual groups, femoral or radial artery access is manually achieved using the modified Seldinger technique. A 5-Fr angiographic catheter is connected to a Y-valve, paired with a sustained high-pressure heparinized saline infusion system. Subsequently, a 0.035-inch guidewire is inserted into the Y-valve. Preoperative assembly procedures for catheters and guidewires are identical between the two groups. The key difference lies in the robotic-assisted group: the entire robotic platform is covered with sterile transparent drapes, and robotic cassettes are assembled. The catheter, Y-valve, and guidewire are then loaded into the respective robotic cassettes, followed by manual insertion of the angiographic catheter into the arterial sheath. During the procedure, physicians in the robotic-assisted group seated in a console room and controlled fluoroscopy and operated the robot via foot treadles and two control sticks. Movements of the catheter and guidewire manipulators on the robotic side are synchronized in real time with those of the control sticks, ensuring seamless and precise operation (Figure 3). The physician receives continuous video feedback and communicates with the patients via a microphone. For the control group, physicians perform the entire angiography process manually using conventional techniques. Physicians in both groups are required to complete selective catheterization of bilateral ICAs and bilateral VAs. Catheter exchanges due to anatomical considerations are permitted in both groups during the procedure. Based on the physician’ s professional judgment, manual intervention may be initiated in the event of an emergency or if the robot is unable to reach the target vessels. However, any conversion from robotic to manual intervention is documented and classified as a technical failure.

**Figure 3.**
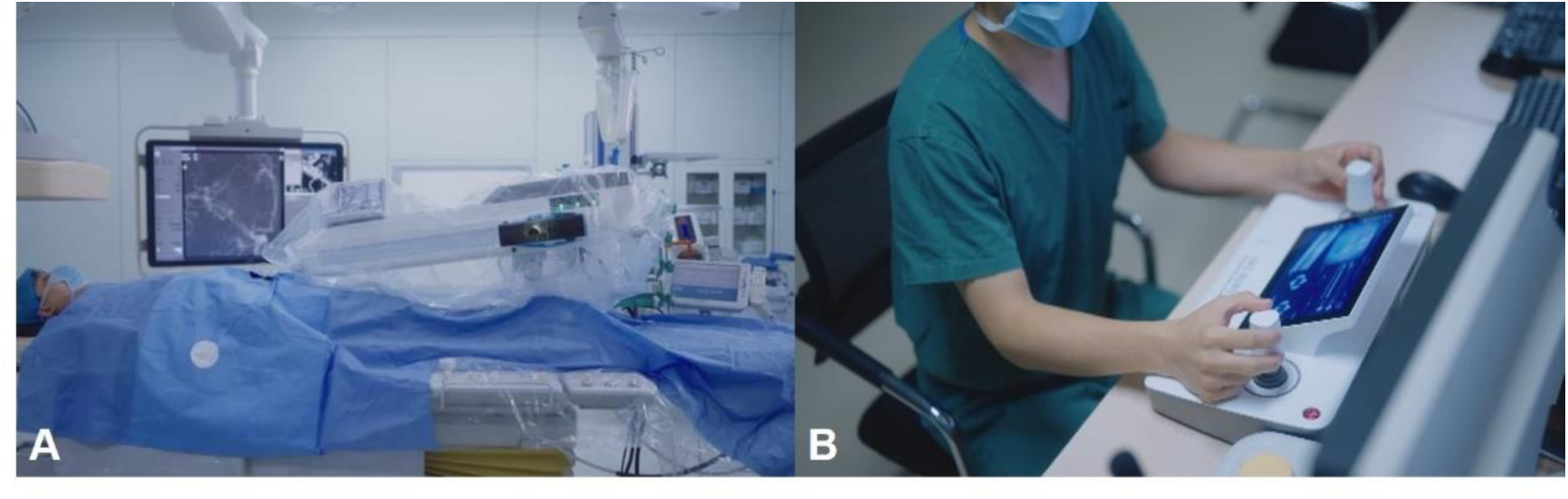
Working status of the YDHB-NS01 Ver 2.0 robotic-assisted system in real-world operating room settings. (A) The robot works in the operating room. (B) The neurointerventionalist controls the robot outside of the operating room (in the control device room).

### Outcomes and definitions

The primary outcome is the clinical success rate, defined as the successful completion of cerebral angiography by the neurointerventional physician (either via the robotic-assisted system or manual technique). Success is confirmed by two criteria: (1) clear visualization of all pre-specified target vessels (without significant imaging artifacts) and (2) no procedure-related serious adverse events (SAEs) occurring intraoperatively or within 24 hours post-procedure.

The secondary outcomes include: technical failure rate, overall procedural time, pre-puncture setup time, mean catheterization time per target vessel, DSA fluoroscopy time, participant radiation dose, primary operator radiation dose, contrast agent volume, and instrument performance evaluation. Definitions of key secondary outcomes are as follows:

Technical failure rate: Defined as any conversion from robotic-assisted to manual operation, or inability to complete the procedure due to robotic system malfunctions (e.g., navigation errors, mechanical arm failure).

Overall procedural time: The duration from the patient being positioned on the operating table to the completion of the procedure (including arterial sheath removal). Pre-puncture setup time: The total time elapsed for patient positioning, disinfection, sterile draping, robotic system assembly (robotic-assisted group only), and pre-procedural device preparation (e.g., catheter/guidewire loading).

DSA fluoroscopy time: Total duration of fluoroscopy during DSA, recorded directly by the DSA system.

Participant radiation dose: The maximum skin radiation dose absorbed by the participant, measured via the DSA system’ s built-in dosimetry module.

Primary operator radiation dose: Cumulative ionizing radiation dose absorbed by the primary operator, measured using a pocket-sized personal dosimeter worn during the procedure.

Contrast agent volume: Total volume of iodinated contrast agent administered during the entire angiographic procedure.

Instrument performance evaluation: Subjective assessment of the robotic system’ s performance (e.g., navigational precision, ease of operation, stability) by the operating physician, completed via a standardized 10-point Likert scale questionnaire immediately post-procedure.

The safety endpoints include:

Incidence of perioperative vascular injuries (intraoperative or within 24 hours post-procedure), including but not limited to vascular perforation, dissection, and pseudoaneurysm;

Occurrence of adverse events (AEs) and SAEs, as defined by the International Council for Harmonisation (ICH) Good Clinical Practice (GCP) guidelines;

Robotic device malfunctions (documented with type, timing, and impact on the procedure);

New asymptomatic cerebral infarctions, assessed via postoperative brain magnetic resonance imaging (MRI) with diffusion-weighted imaging (DWI) (performed within 24 hours post-procedure for all participants).

### Follow-up schedule

All participants will undergo follow-up assessments conducted by on-site neurosurgeons and/or neurointerventionalists at baseline, on the day of the DSA procedure, and at hospital discharge. All patients are required to undergo brain MRI within 24 hours post-DSA to evaluate for new asymptomatic cerebral infarctions. Telephone follow-up will be performed 7 days after discharge. During the follow-up assessment, the Modified Rankin Scale (mRS) score, potential AEs/SAEs, and study endpoints will be reviewed by experienced neurosurgeons and/or neurointerventionalists blinded to the group assignment. Table 2 outlines the comprehensive study assessment schedule.

**Table 2.** ERASE trial assessment schedule.

| Procedure/ Assessment | Time Frame |  |  |  |
| --- | --- | --- | --- | --- |
|  | Basel<br>ine | End of<br>surgery | 24 hours<br>post-sur<br>gery | 7 days<br>post-disc<br>harge |
| Informed Consent | √ | / | / | / |
| Demographic Data | √ | / | / | / |
| Comorbidities and medical history | √ | / | / | / |
| Modified Rankin Scale (mRS) | √ | √ | √ | √ |
| Physical examination | √ | / | √ | / |
| Routine laboratory tests | √ | / | √ | / |
| Electrocardiography | √ | / | / | / |
| Brain MRI | √ | / | √ | / |
| American Society of Anesthesiologists (ASA)<br>Physical Status Classification | √ | / | / | / |
| Verify inclusion/exclusion criteria | √ | / | / | / |
| Randomization | √ | / | / | / |
| Clinical Success Rate | / | √ | / | / |
| Super-selective Technique Success Rate | / | √ | / | / |
| Super-selective Catheterization Time per Target<br>Vessel | / | √ | / | / |
| Pre-puncture setup time | / | √ | / | / |
| DSA Fluoroscopy Time | / | √ | / | / |
| Overall Procedural Time | / | √ | / | / |
| Participant Radiation Dose | / | √ | / | / |
| Primary Operator Radiation Dose | / | √ | / | / |
| Contrast Agent Volume | / | √ | / | / |
| Instrument Performance Evaluation | / | √ | / | / |
| Assessment of Adverse events (AEs) and (SAEs) | / | √ | √ | √ |

ERASE, Endovascular Robotic Assisted SystEm for Cerebral Angiography; MRI, magnetic resonance imaging.

### Data Collection and Management

Individual patient data will be documented in standardized case report forms (CRFs) and entered into a web-based electronic data capture (EDC) system. Data transmission between the user browser and the EDC database will utilize a secure, encrypted connection, and database access will be restricted via role-based password protection. A designated clinical research monitor will regularly verify the completeness, accuracy, and consistency of CRF data against source documents.

### Data Monitoring Committee

An independent Data Monitoring Committee (DMC, unaffiliated with the study investigators or sponsor) will conduct periodic reviews of trial progress and patient safety. The DMC will assess the need for protocol amendments, interim analyses, or early trial termination. Any proposed modifications to the inclusion/exclusion criteria, outcome measures, or other key protocol elements will be formally documented in a protocol amendment. This revised protocol will then be submitted to the IRB of each participating center for reapproval before implementation.

### Sample Size Calculation

The primary hypothesis is that the clinical success rate of the robotic-assisted group is non-inferior to that of the control group. Based on prior clinical data and published literature, the clinical success rate of manual cerebral angiography is estimated to be 95–98%. Using a non-inferiority margin of 8% (i.e., the maximum acceptable difference in success rate between the robotic-assisted and manual groups), a two-sided significance level (α) of 0.05, and a statistical power (1−β) of 80%, sample size calculation was performed using PASS 15.0 software. This yielded a required total sample size of 386 participants. Accounting for an anticipated 15% withdrawal or loss-to-follow-up rate, the final enrollment target is set at 450 participants to ensure sufficient statistical power for the primary endpoint analysis.

## Statistical analyses

Statistical analyses will be performed using SPSS 26.0 software (IBM Corp., Armonk, NY, USA). Continuous variables will be summarized as mean ± standard deviation (SD), median, lower quartile (Q1), and upper quartile (Q3). Normality of distribution will be assessed using the Shapiro-Wilk test. Categorical variables will be presented as frequencies and percentages (n, %), with incidence rates calculated where appropriate.

The primary endpoint (clinical success rate) will be analyzed using a two-sample test for non-inferiority based on the asymptotic Z-test. Non-inferiority will be confirmed if the lower bound of the two-sided 95% confidence interval for the between-group difference (robotic-assisted group − control group) exceeds −8%. Normally distributed continuous variables will be compared between groups using the student’ s t-test; non-normally distributed continuous variables will be analyzed using the Wilcoxon rank-sum test. Categorical variables will be compared using the chi-square test or Fisher’ s exact test. All statistical tests will be two-sided, and a P value < 0.05 will be considered statistically significant. Analyses will be performed based on the intention-to-treat principle, with the per-protocol set used for sensitivity analysis to verify the robustness of results.

## Discussion

Robots have increasingly established themselves as indispensable tools across a broad spectrum of medical procedures with substantial promise and clinical necessity in interventional surgery. During interventional procedures, interventionalists typically stand for prolonged periods wearing heavy personal protective equipment, which places sustained physical strain on the musculoskeletal system and contributes to cervical and lumbar injuries ^12^. Diagnostic cerebral angiography constitutes a major component of neurointerventional practice, and robotic systems offer a critical advantage by reducing operator radiation exposure and other occupation-related hazards ^17^. Prior studies have linked spinal disorders to annual procedural volume and years of clinical practice ^18^, an issue where robotic assistance provides benefits by alleviating physical demands and mitigating operator fatigue. Additionally, interventionalists can remotely operate the procedure while seated comfortably, eliminating the need for lead aprons, thereby minimizing back discomfort and reducing the risk of orthopedic injuries ^7^.

Beyond these benefits, the advantages of robotic-assisted interventional systems extend further. Since 2012, the CorPath robotic platform (Corindus, Waltham, Massachusetts, USA) has been utilized for percutaneous coronary interventions ^12^ ^19^. Key insights from its use in this setting highlight that the primary benefits of robotic assistance include enhanced procedural precision, improved technical accuracy, and reduced radiation exposure during fluoroscopic imaging ^18^. However, a critical limitation of robotic-assisted systems is their reliance on visual cues, such as subtle changes in device shape or motion to detect vascular obstacles and friction. This visual feedback may be insufficient to offset the loss of tactile feedback that is inherent to manual interventional procedures. While subsequent studies on neurointerventional robotic systems have similarly emphasized their potential to improve surgical precision and safety ^14–16^, these purported advantages necessitate validation through more objective, evidence-based metrics.

Beyond established risk factors such as prolonged procedural duration and advanced age, our team hypothesizes that certain intraprocedural interventional techniques are also closely intertwined with this outcome. Specifically, repeated guidewire exchanges, frequent switching of high-pressure heparinized saline infusions, and unrecognized microbubbles entering the Y-valve may all generate microemboli during the procedure, contributing to positive postoperative DWI findings. To address these concerns, the YDHB-NS01 Ver 2.0 robotic-assisted system incorporates targeted design enhancements: (1) a specialized one-way valve mechanism that obviates the need for repeated switching of high-pressure heparinized saline infusions; (2) the ability to facilitate angiography without complete guidewire withdrawal; and (3) a dedicated monitoring camera at the Y-valve to alert operators to microscopic bubbles that might otherwise go undetected. These integrated optimizations collectively mitigate the risk of the aforementioned intraprocedural complications, reinforcing the system’ s safety advantage.

In the ERASE trial, we advance the evidence base by demonstrating these direct clinical benefits for patients undergoing cerebral angiography. A key strength of our study is the integration of postoperative DWI findings comparison between the two groups into the safety endpoint assessment. This design aims to yield higher-quality evidence for the safety profile of robotic-assisted neurointerventional procedures. Prior studies have reported that approximately 20% of patients develop positive DWI lesions following neurointerventional surgery ^20–24^. Beyond established risk factors such as prolonged procedural duration and advanced age, our team hypothesizes that certain intraprocedural interventional techniques are also closely intertwined with this outcome. Specifically, repeated guidewire exchanges, frequent switching of high-pressure heparinized saline infusions, and unrecognized microbubbles entering the Y-valve may all generate microemboli during the procedure ^25^, contributing to positive postoperative DWI findings. To address these concerns, the YDHB-NS01 Ver 2.0 robotic-assisted system incorporates targeted design enhancements: (1) a specialized one-way valve mechanism that obviates the need for repeated switching of high-pressure heparinized saline infusions; (2) the ability to facilitate angiography without complete guidewire withdrawal; and (3) a dedicated monitoring camera at the Y-valve to alert operators to microscopic bubbles that might otherwise go undetected. These integrated optimizations collectively mitigate the risk of the aforementioned intraprocedural complications, reinforcing the system’ s safety advantage.

Additionally, we have implemented a speed limitation for catheter and guidewire advancement via the robotic delivery system, it was designed to prevent vascular injury associated with overly rapid manipulation and mitigate the risk of air bubbles entering the Y-valve or catheters during guidewire withdrawal. Manual cerebral angiography is analogous to a driver operating a vehicle: an experienced operator may perform manipulations beyond ‘safe thresholds’, much like a driver exceeding an 80 km/h speed limit to 100 km/h without incident. Yet this inherent safety risk is magnified by operator fatigue, distractions, or human variability. By contrast, a robotic-assisted interventional system operates like a vehicle’ s cruise control: it adheres strictly to preset speed limits, functioning without emotional bias or fatigue, and delivering consistent, unwavering performance. This aligns closely with the core objective of our study and what we aim to validate through the ERASE trial: standardizing physician-performed interventional workflows to minimize patient harm stemming from interoperator technical variability.

## Summary and Conclusions

The ERASE trial is a randomized controlled study designed to evaluate the efficacy and safety of the YDHB-NS01 Ver 2.0 endovascular robotic-assisted system in patients scheduled for cerebral angiography. The primary efficacy outcome is the clinical success rate, while the primary safety outcome is the incidence of perioperative and postoperative complications and device malfunctions. Enrollment of the first participant in ERASE commenced on September 1, 2025. As of December 1, 2025, 80 participants had been enrolled. The trial is anticipated to be completed by late May 2027. The present manuscript details the rationale and design of the ERASE trial.

## Acknowledgements

We thank all the participants in the ERASE program for their hard work.

## Disclosures

### Contributors

SG, YL and YZ conceptualized and designed the initial protocol. JJ, XX and CL amended the initial protocol. YZ, SG and JJ drafted the manuscript. YZ, QL, JC, AL, XZ, FW, XZ, JL, YJ, XL, YG, XM, WY, WG, YS and SG contributed to the acquisition of data. All authors have read and approved the final version of the manuscript to be published.

Competing interests None declared.

### Ethics approval Statement

This study involves human participants and all protocol modifications were submitted and approved by the institutional review board at Peking Union Medical College Hospital (IRB approval number: I-24PJ1748) and all participating centers. The study is conducted in accordance with Good Clinical Practice and the Declaration of Helsinki. All patients or their legal representatives must provide written consent.

### Data availability statement

All data are available to researchers on request for purposes of reproducing the results or replicating the procedure by directly contacting the corresponding author.

## Abbreviations

ERASE: Endovascular Robotic-Assisted SystEm for Cerebral Angiography
RCT: randomized controlled trial
MRI: magnetic resonance imaging
DSA: digital subtraction angiography
DWI: diffusion-weighted imaging
IRB: institutional review board
ICA: internal carotid artery
IWRS: interactive web response system
VA: vertebral artery
AE: adverse event
SAE: serious adverse event
ICH: International Council for Harmonisation
GCP: Good Clinical Practice
CRF: case report form
EDC: electronic data capture
DMC: Data Monitoring Committee
SD: standard deviation

## References

1. Thilak S, Brown P, Whitehouse T, et al. Diagnosis and management of subarachnoid haemorrhage. Nat Commun 2024;15(1):1850. doi: 10.1038/s41467-024-46015-2 [published Online First: 2024/03/01]

2. Nogueira RG, Jovin TG, Liu X, et al. Endovascular therapy for acute vertebrobasilar occlusion (VERITAS): a systematic review and individual patient data meta-analysis. Lancet 2025;405(10472):61–69. doi: 10.1016/s0140-6736(24)01820-8 [published Online First: 2024/12/15]

3. Han H, Chen Y, Ma L, et al. Interventional Treatment vs Conservative Management of Unruptured Brain Arteriovenous Malformations. JAMA Netw Open 2025;8(11):e2543408. doi: 10.1001/jamanetworkopen.2025.43408 [published Online First: 2025/11/13]

4. Liu J, Ni W, Zuo Q, et al. Middle Meningeal Artery Embolization for Nonacute Subdural Hematoma. N Engl J Med 2024;391(20):1901–12. doi: 10.1056/NEJMoa2401201 [published Online First: 2024/11/20]

5. Nguyen TN, Abdalkader M, Fischer U, et al. Endovascular management of acute stroke. Lancet 2024;404(10459):1265–78. doi: 10.1016/s0140-6736(24)01410-7 [published Online First: 2024/09/29]

6. Sun X, Deng Y, Zhang Y, et al. Balloon Angioplasty vs Medical Management for Intracranial Artery Stenosis: The BASIS Randomized Clinical Trial. Jama 2024;332(13):1059–69. doi: 10.1001/jama.2024.12829 [published Online First: 2024/09/05]

7. Sajja KC, Sweid A, Al Saiegh F, et al. Endovascular robotic: feasibility and proof of principle for diagnostic cerebral angiography and carotid artery stenting. J Neurointerv Surg 2020;12(4):345–49. doi: 10.1136/neurintsurg-2019-015763 [published Online First: 2020/03/03]

8. Snelling BM, Sur S, Shah SS, et al. Transradial cerebral angiography: techniques and outcomes. J Neurointerv Surg 2018;10(9):874–81. doi: 10.1136/neurintsurg-2017-013584 [published Online First: 2018/01/10]

9. Pelz DM, Fox AJ, Vinuela F. Digital subtraction angiography: current clinical applications. Stroke 1985;16(3):528–36. doi: 10.1161/01.str.16.3.528 [published Online First: 1985/05/01]

10. Alakbarzade V, Pereira AC. Cerebral catheter angiography and its complications. Pract Neurol 2018;18(5):393–98. doi: 10.1136/practneurol-2018-001986 [published Online First: 2018/07/20]

11. Henkes H, Khanafer A, Cimpoca A. Safety of Cerebral Angiography. Cardiovasc Intervent Radiol 2023;46(7):929–30. doi: 10.1007/s00270-023-03471-5 [published Online First: 2023/06/01]

12. Maor E, Eleid MF, Gulati R, et al. Current and Future Use of Robotic Devices to Perform Percutaneous Coronary Interventions: A Review. J Am Heart Assoc 2017;6(7) doi: 10.1161/jaha.117.006239 [published Online First: 2017/07/26]

13. Crinnion W, Jackson B, Sood A, et al. Robotics in neurointerventional surgery: a systematic review of the literature. J Neurointerv Surg 2022;14(6):539–45. doi: 10.1136/neurintsurg-2021-018096 [published Online First: 2021/11/21]

14. Liu H, Li C, Ren S, et al. Efficacy and safety of a neurointerventional operation robotic assistance system in cerebral angiography. Stroke Vasc Neurol 2024;9(3):243–51. doi: 10.1136/svn-2022-002260 [published Online First: 2023/08/24]

15. Zhang Y, Xu H, Sun J, et al. Safety and efficacy of cerebral robot assisted angiography: randomized comparison of robotic versus manual procedures. J Neurointerv Surg 2025 doi: 10.1136/jnis-2025-023412 [published Online First: 2025/07/10]

16. Beaman C, Gautam A, Peterson C, et al. Robotic Diagnostic Cerebral Angiography: A Multicenter Experience of 113 Patients. J Neurointerv Surg 2024;16(7):726–30. doi: 10.1136/jnis-2023-020448 [published Online First: 2023/07/20]

17. Ross AM, Segal J, Borenstein D, et al. Prevalence of spinal disc disease among interventional cardiologists. Am J Cardiol 1997;79(1):68–70. doi: 10.1016/s0002-9149(96)00678-9 [published Online First: 1997/01/01]

18. Goldstein JA, Balter S, Cowley M, et al. Occupational hazards of interventional cardiologists: prevalence of orthopedic health problems in contemporary practice. Catheter Cardiovasc Interv 2004;63(4):407–11. doi: 10.1002/ccd.20201 [published Online First: 2004/11/24]

19. Weisz G, Metzger DC, Caputo RP, et al. Safety and feasibility of robotic percutaneous coronary intervention: PRECISE (Percutaneous Robotically-Enhanced Coronary Intervention) Study. J Am Coll Cardiol 2013;61(15):1596–600. doi: 10.1016/j.jacc.2012.12.045 [published Online First: 2013/03/19]

20. McDonnell CO, Fearn SJ, Baker SR, et al. Value of diffusion-weighted MRI during carotid angioplasty and stenting. Eur J Vasc Endovasc Surg 2006;32(1):46–50. doi: 10.1016/j.ejvs.2005.12.026 [published Online First: 2006/03/21]

21. Carraro do Nascimento V, de Villiers L, Hughes I, et al. Transradial versus transfemoral arterial approach for cerebral angiography and the frequency of embolic events on diffusion weighted MRI. J Neurointerv Surg 2023;15(7):723–27. doi: 10.1136/jnis-2022-019009 [published Online First: 2022/07/23]

22. Carraro do Nascimento V, de Villiers L, Dhillon PS, et al. Transradial versus transfemoral access for diagnostic cerebral angiography: frequency of acute MRI findings in 500 consecutive patients at a single center. J Neurointerv Surg 2025;17(2):181–85. doi: 10.1136/jnis-2024-021472 [published Online First: 2024/03/20]

23. Habtezghi AB, Ghozy S, Bilgin C, et al. DWI-Detected Ischemic Lesions after Endovascular Treatment for Cerebral Aneurysms: An Updated Systematic Review and Meta-analysis. AJNR Am J Neuroradiol 2023;44(11):1256–61. doi: 10.3174/ajnr.A8024 [published Online First: 2023/10/13]

24. Bendszus M, Koltzenburg M, Burger R, et al. Silent embolism in diagnostic cerebral angiography and neurointerventional procedures: a prospective study. Lancet 1999;354(9190):1594–7. doi: 10.1016/s0140-6736(99)07083-x [published Online First: 1999/11/24]

25. Cruz AS, Khattar NK, Weiner GM, et al. Preventing air microembolism in cerebral angiography: a JNIS fellow’s perspective. J Neurointerv Surg 2024;16(4):331–32. doi: 10.1136/jnis-2024-021653 [published Online First: 2024/03/15]

